# Mixture of organic pollutants is associated with cognitive aging

**DOI:** 10.1101/2025.10.17.25338236

**Authors:** Vrinda Kalia, Katherine E. Manz, Jaime Benavides, Suhang Song, Brandi L. Vollmer, Beizhan Yan, Jeff Goldsmith, Kurt D. Pennell, Yaakov Stern, Marianthi-Anna Kioumourtzoglou, Gary W. Miller, Christian Habeck, Yian Gu

**Author notes:** Department of Epidemiology, Brown University School of Public Health, Providence, RI 02903. a) Department of Epidemiology, Brown University School of Public Health, Providence, RI 02903, b) Institute at Brown University for Environment and Society, Providence, RI 02912. Corresponding author: Vrinda Kalia, PhD, MPH 650 W 168^th^ street, Room 1618 New York, NY 10032.

## Abstract

**INTRODUCTION:** Environmental organic pollutants impact brain function and cognitive aging, but the effects of real-world complex mixtures of these pollutants is unexplored.

**METHODS:** Using data collected at two time points from 170 cognitively normal adults, we used hierarchical Bayesian kernel machine regression to examine the association between joint exposure to 49 organic pollutants and latent variables, derived from neuropsychological tests, that capture key aspects of cognitive aging.

**RESULTS:** We observed a non-linear, inverted U-shaped relationship between the pollutant mixture and the global cognitive score. Polychlorinated biphenyls (PCBs) were the most important pollutant group in the mixture followed by industrial-use pollutants.

**DISCUSSION:** Exposure to a mixture of organic pollutants was associated with poor cognitive aging. Even though many of these pollutants, like PCBs, have been banned for decades, they persist in our environment. Strategies to reduce exposure to these organic pollutants are needed to minimize their impact on cognitive aging.

## 1. Background

A person’s cognitive abilities have profound impacts on their day-to-day activities, directly influencing their human experience and contribution to society. Cognitive abilities are also an important factor for successful health and aging [1]. As lifespans across the world are increasing, there is an increased need to understand factors that affect cognitive aging, i.e., decline in cognitive performance over age. Several studies point to a role for the environment in cognitive performance over adulthood. Identifying modifiable risk factors, such as environmental pollutants, is an important step toward alleviating the burden of poor cognitive aging in an aging population.

The environment has been shown to affect cognitive function in different ways. Cognitive performance can be enhanced by improved ambient environment in buildings [2,3], while exposure to certain chemical pollutants has been associated with poorer cognitive performance. For example, exposure to air pollution has been associated with poor cognitive performance in older adults across many countries [4–8], as has exposure to heavy metals, including arsenic, lead, cadmium, and rubidium [9–11]. Fewer studies have explored the relationship of organic pollutant exposures, like polychlorinated biphenyls (PCBs) or organochlorine pesticides [12] with cognitive function and only a handful have applied a mixtures approach [13–15], which considers exposure to multiple chemicals simultaneously in health analyses. Furthermore, no study to date has investigated the relationship between organic pollutant exposure and aspects of cognitive function most relevant to cognitive aging. By identifying environmental exposures that influence cognitive aging, we can propose targeted interventions to reduce exposure to adverse agents through policy or individual action with the intention to decelerate cognitive aging. Additionally, by uncovering specific sources of pollutants that are the most consequential in this relationship, we can identify interventions with the greatest benefit.

Salthouse [16] argues that many neuropsychological or cognitive variables are related to each other and that by treating them as independent variables when considering the effect of specific variables, the estimated magnitudes of associations may be misleading [17]. Using large neuropsychological test batteries conducted over a life course, Salthouse and colleagues applied a multivariate approach and discovered four latent variables that capture most of the variance in age-related cognitive decline [16–18]. These include the cognitive domains: episodic memory, fluid reasoning, processing speed, and vocabulary. We leveraged these previously constructed distinct cognitive domain scores in two ongoing studies: the cognitive reserve (CR) study and the reference ability neural network study (RANN) [19,20], to identify environmental organic pollutants that are associated with cognitive aging. By considering the derived cognitive domain, instead of each specific neuropsychological variable, we take a more efficient approach [17,19] at studying the influence of these organic pollutants on cognitive aging.

Using banked serum samples collected at two time points from 170 participants of the CR/RANN study, we determined the relationship between exposure to organic pollutants and the cognitive domains. We applied hierarchical Bayesian kernel machine regression (BKMR), a mixtures analysis approach that estimates a flexible multivariable exposure-response function, allowing us to investigate potential non-linear relationships between the exposures and health outcome. It also incorporates hierarchical variable selection, providing a way to identify which group(s), and individual component(s) within a group, are most likely contributing to the estimated health effect [21]. Using hierarchical BKMR, we examined 1) whether the organic pollutants measured in serum had an overall mixture effect, 2) whether pollutants from a specific source were particularly important to the relationship, and 3) which specific pollutants played an important role in the relationship within each group. This study is the first to provide evidence of a relationship between organic pollutant exposure and cognitive aging.

## 2. Methods

### 2.1 The participants

The participants were part of two ongoing studies at the Columbia University Irving Medical Center: the CR and the RANN study [19,20]. The participants were between the ages of 20 – 80 years, had to be native English speakers, strongly right-handed, and have at least fourth grade reading level. They were screened for MRI contraindications, hearing or visual impairment, medical or psychiatric conditions that could affect cognition, and dementia or mild cognitive impairment in older (age > 55 years) participants. Additionally, participants were required to perform within age-adjusted normal limits on a list-learning test and have no complaints on a functional impairment questionnaire [19]. Participants also provided blood samples that were used for genotyping, and serum was banked for future analysis. The neuropsychological test data, neuroimaging data, and blood samples were also collected at a follow-up visit, ∼5 years after the initial assessment. From more than 500 participants in these studies, 170 unique participants were included in this analysis, 164 with data collected both timepoints, and six with data only at one time point. There was no difference in the participants included in this study compared to those excluded (Table S1). The study was approved by Columbia University’s Institutional Review Board and all participants provided written consent.

### 2.2 Assessment of cognitive function

Twelve measures from a battery of neuropsychological tests were selected to assess functioning in four cognitive domains. Based on a principal axis factor analysis, the composite scores for each of the four cognitive domains were determined by specific sets of tasks. These tasks included: 1) Wechsler Adult Intelligence Scale (WAIS-III) Block design task, WAIS-III Letter–Number Sequencing test, and WAIS III Progressive Matrices for fluid reasoning; 2) WAIS-III Digit Symbol Subtest, Part A of the Trail Making Test, and the Stroop Color Naming tests for processing speed; 3) the vocabulary subtest from WAIS-R, the Wechsler Test of Adult Reading (WTAR), and the American National Adult Reading Test (AMNART) for vocabulary; and 4) three sub-scores of the Selective Reminding Task (SRT): Long-Term Storage sub-score (SRT LTS), Continuous Long-Term Retrieval (SRT CLRT), and Last Trial (SRT Last) for episodic memory. Z-scores were computed for participants, on each summary score, based on the overall means and standard deviations. A higher score indicated better cognitive performance. Along with a score on each cognitive domain, a composite global score was also generated as the average of z-scores on all four domains. We considered the global score as the primary outcome of interest and secondarily explored the four cognitive domains to see whether specific domains were more likely to be impacted by pollutants.

### 2.3 Gas Chromatography coupled high resolution mass spectrometry (GC-HRMS) assay

GC-HRMS was used to measure levels of organic pollutants in 170 participants using serum samples collected at two points (334 total samples). The samples were treated using a QuEChERS extraction [22]. Briefly, samples were sonicated in 1 mL hexane:acetone: dichloromethane and transferred to a 2 mL QuEChERS tube containing 150 mg MgSO_4_ and 50 mg C18. The supernatant was transferred to a new vial and the QuEChERS tube was washed with 0.5 mL hexane:acetone:dichloromethane twice. The extract was evaporated under nitrogen to a final volume of 200-300 µL hexane using an Organomation 30 position Multivap Nitrogen Evaporator (Organomation Associates Inc.) and transferred to a glass autosampler vial. The samples were analyzed on 2 analytical columns: one column was used for furans, dioxins, and brominated diphenyl ethers (PBEs) and the second column was used to analyze PCBs, pesticides, and other nonbrominated flame retardants. Sample extracts were analyzed using a high-resolution Thermo Q Exactive Orbitrap MS equipped with a Thermo Trace 1300 GC and a TriPlus RSH Autosampler. Quantification was performed using an 8-point calibration curve using serial dilution of calibration standards in hexane (0.025 – 15 µg/L). A total of 125 pollutants were quantified and included a wide range of persistent organic pollutants: PCBs, organochlorine pesticides, polyaromatic hydrocarbons (PAHs), dioxins, and industrial solvents. The limits of quantification for each chemical can be found in Table S2. These pollutants were chosen based on chemical standards availability and choosing a reasonable number of chemicals that could be mixed to create chemical standards for analysis.

### 2.4 Covariate selection

Covariates of interest included age (in years), sex (male/female), race (White/Black/other), education (in years), analytical batch (batch to which a sample belonged in the GC-HRMS assay), and APOE-ε4 allele status (one or more ε4 allele vs. no ε4). These covariates were chosen a priori based on previous studies [23–25]. We did not have access to variables to inform social economic status (SES) therefore we used education as a proxy for SES.

### 2.5 Exposure data processing

We excluded from analyses pollutants detected in less than 50% of the samples (Table S2). We also filtered pollutants using the coefficient of quartile variation (CQV), to allow for enough variability—and thus information. We retained those pollutants with a CQV ≥ 0.6 at each individual time point and averaged across the two time points. Forty-nine chemicals met these inclusion criteria. The pollutant concentrations were log_2_-transformed and scaled before statistical analysis.

### 2.6 Chemical source assignment

To determine if pollutants from a particular expected environmental source were associated with cognitive aging, we assigned each pollutant to one possible source. These were assigned based on a review of literature, EPA pesticide documentation and register, and expert knowledge of two analytical chemists. In cases where more than one possible source was identified, the source most likely to lead to detection in the blood stream was selected. The possible sources were: 1) food: this included PAHs and dioxins that are most likely to be found in serum by exposure through food (p = 4 pollutants); 2) indoor dust: this included pollutants that have been found in indoor dust, including pollutants found in personal care and other household products [26–29] (p = 10); 3) farming/green space: included pollutants used as pesticides in farms and on urban green spaces (p = 6); 4) industrial use: pollutants used in industrial processes (p = 5); 5) legacy: pollutants no longer in production but postulated to be found in the blood stream through similar routes of exposure (p = 12); 6) legacy PCBs: PCBs no longer in production but assumed to come from the same route of exposure (p = 10); and 7) pollutants that did not fall under these categories were grouped under “other” (p = 2) (Figure 1, Table S3).

**Figure 1.**
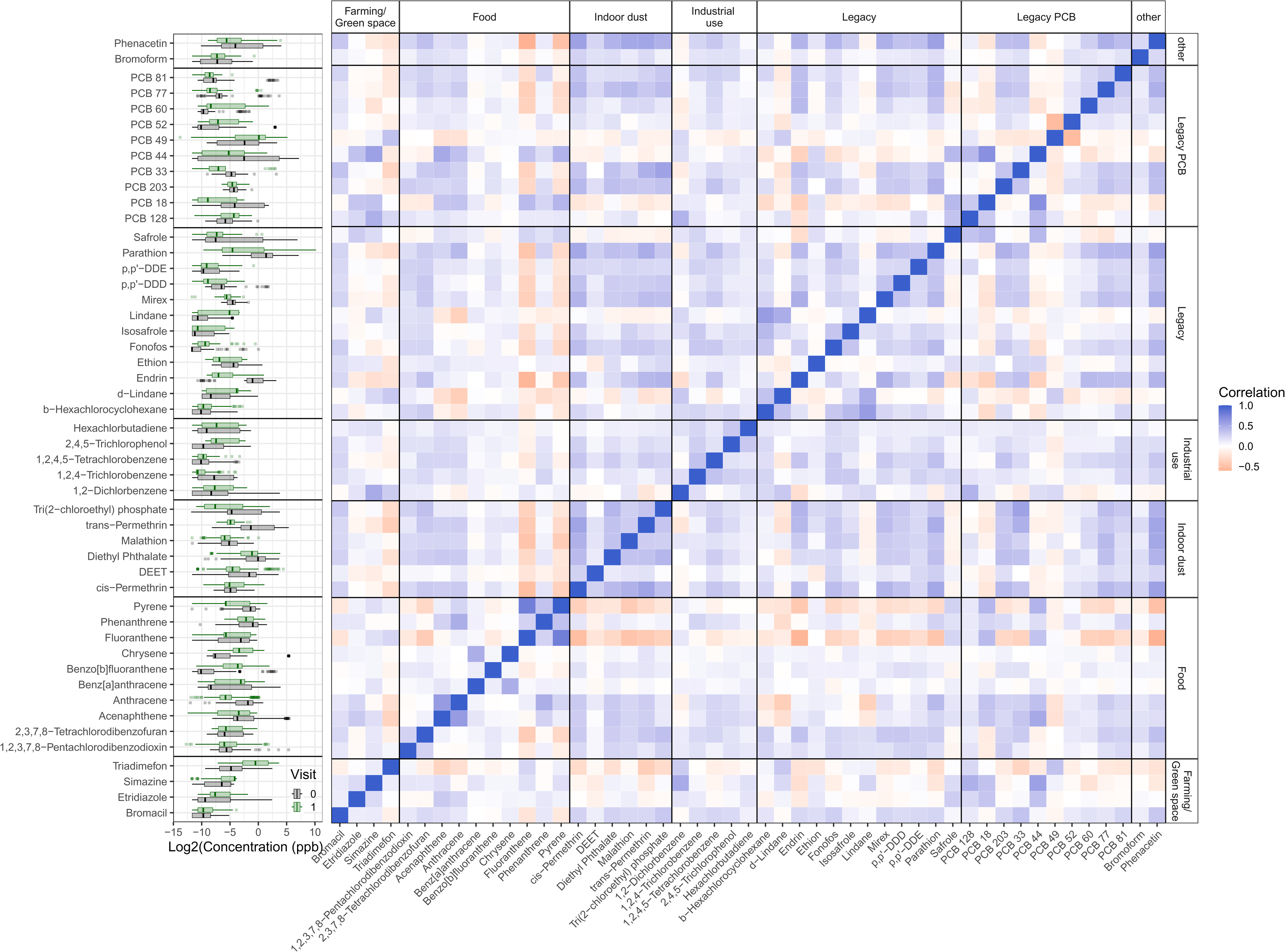
Pollutant distribution and correlation. The distribution of Log2 transformed chemical pollutant concentrations shown in box plots separately for the two visits (grey for the baseline and green for the follow up visit). The heatmap shows the spearman correlation coefficients between all the pollutants, faceted by the assigned source group. The darker the blue color, the greater the positive correlation while the darker the orange, the smaller the negative correlation.

### 2.7 Single-pollutant models

We used generalized estimating equations (GEE) to determine the relationship between each pollutant and each cognitive domain and the global score, adjusted for covariates described in section 2.4 using an exchangeable correlation structure and robust standard errors to account for the repeated outcome observations within each participant. We used a false discovery rate of 5% to control for multiple hypothesis testing. We used the *gee()* function in the R package “gee” (version 4.13.29) to run the models.

### 2.8 Hierarchical Bayesian Kernel Machine Regression (BKMR)

We used hierarchical BKMR [21] to consider the 49 pollutants as a mixture in the health analysis. Data from each timepoint were included and the exposures and outcome were analyzed as repeated measures. We used BKMR since it allows for 1) flexible (i.e., non-linear) relationships between highly correlated exposures and outcome, 2) incorporation of *a priori* assigned chemical grouping, 3) multiple observations per person by the inclusion of subject-level random intercepts, and 4) covariate adjustment [21]. Operationally, the kernel machine regression may be expressed as a mixed-effects model, and in a Bayesian context, non-informative prior distributions are placed on all unknown parameters [30]. We used the hierarchical variable selection approach with 60,000 iterations by a Markov chain Monte Carlo (MCMC) algorithm using the *kmbayes()* function in the R package “bkmr” [30] (version 0.2.2). The overall mixture effect can be operationalized as the combined effect of the pollutants in the mixture, estimated using a non-parametric Gaussian kernel function within a Bayesian regression model [31,32]. The kernel machine representation regularizes the high-dimensional exposure-response function and induces a correlation of health outcome among individuals with similar exposure profiles, i.e., it assumes that two individuals with similar exposure values have similar health risks, given covariates.

To determine if pollutants from a certain source were more important in explaining the relationship between exposure and response, we performed a hierarchical analysis. This provides group (pollutant source) and within-group posterior inclusion probabilities (PIPs), giving an estimate of the number of times each group and pollutant were selected to be included in the exposure-response function across iterations. A higher PIP suggests a variable is a likely predictor of the health outcome. Variable selection is performed using an augmented Gaussian kernel function with a spike-and-slab prior framework, a two-part prior, for the auxiliary parameters. This approach includes an indicator that can be interpreted as the PIP for a mixture component [21,33].

We ran separate models for the global score, and secondarily each cognitive domain, as the outcome and all 49 pollutants as the mixture. We present the estimated relationships between each outcome and pollutant exposure while fixing the other pollutants at their median exposure level. We also present the estimated overall mixture effect when considering the estimated relationship between the mixture exposure and outcome when all chemicals were fixed at a particular percentile, ranging from 25^th^ – 75^th^ percentile, with 5% increments, using the median as the reference.

To evaluate BKMR performance, we visually inspected the MCMC trace plots for each outcome investigated, plotting the coefficient for age and residual variance (a^2^) after excluding the burn-in (the first 5,000 iterations).

### 2.9 Statistical software and code availability

All statistical analyses were conducted in R (version 4.4.1). The code used for analysis was reviewed by a co-author and can be found on the first author’s GitHub page: https://github.com/vrindakalia/chemicals_and_reference_abilities

## 3. Results

### 3.1 Study population characteristics

The average age of the participants across both visits was 59 (SD = 16.4) years. There were nearly equal number of males and females (53% women), 68% of the participants were White, 23% were Black, and people of Asian or mixed race, classified as “other”, accounted for 9% of the sample. On average, the participants had 16 years (range: 12-24) of education. About 25% of the participants had at least one APOE-ε4 allele (Table 1).

**Table 1.** Characteristics of participants in the study.

### 3.2 Exposure measurement and source assignment

One hundred and twenty-five pollutants were measured, of which 57 were detected in at least 50% of all the samples (Table S2), and 49 of these had a CQV ≥ 0.6 at each visit and the average of both visits. The mean and median concentrations of these pollutants can be found in Table S3.

### 3.3 Correlations among pollutants

As expected, correlations were generally higher within each group than across groups. Within each group, the strongest positive Spearman correlations were between fluoranthene and pyrene (r = 0.78) in the food group, 1,2,4-trichlorobenzene and 1,2,4,5-tetrachlorobenzene (r = 0.35) in the industrial use group, etridiazole and simazine (r = 0.21) in the farming/ green space group, cis-Permethrin and trans-Permethrin in the indoor dust group (r = 0.54), b-Hexachlorocyclohexane and lindane (r = 0.59) in the legacy group, and PCB 44 and PCB 18 (r = 0.67) in the legacy PCB group. All pollutants in the indoor dust and industrial use group were either positively correlated or not correlated with each other, i.e. none were negatively correlated (range: 0.2 – 0.54, Figure 1, Table S4, Figure S1).

### 3.4 Single-pollutant models

After adjusting for multiple comparisons, none of the pollutants were associated with any of the cognitive domains or the global cognitive score using GEE controlling for the covariates described in section 2.4 (Figure S2).

### 3.5 Mixture analysis of association between pollutants on global cognitive score

We considered the global score as the primary outcome of interest since it is a composite score of the four cognitive domains. Overall, we estimated an inverted U-shaped relationship between the pollutant mixture (overall effect) and global score (Figure 2A). Relative to the reference (all pollutants set to their median values), both increasing and decreasing pollutant percentiles were associated with lower global cognitive scores (Figure 2A). This relationship was statistically significant at lower percentiles. The pollutant groups with the highest posterior probability of inclusion were legacy PCBs and pollutants of industrial use (PIP = 0.74 and 0.66, respectively; Figure 2B). The univariate response curves in Figure 2C, show the relationships and 95% credible intervals between the global score and each pollutant while fixing all the other pollutants at their median level. Within legacy PCBs, we observed the strongest, negative associations with PCB 203 and PCB 60. For all other chemicals, regardless of group membership, the credible intervals included the null. Within groups, we found some suggestive evidence of associations with some of the group pollutants, albeit the direction of these associations was not consistent. Some mixture components (and their direction of association in parentheses) of note include: in the farming/green space group, bromacil (positive) and simazine (negative); in the food group, 1,2,3,7,8-pentachlorodibenzodioxin and anthracene (both negative); in the indoor dust group, cis-permethrin (positive), tri(dichloroethyl)phosphate (positive), and diethyl phthalate (negative); in industrial use group, 1,2-dichlorobenzene (positive); and in the legacy group, p,p-DDD and safrole (both positive).

**Figure 2.**
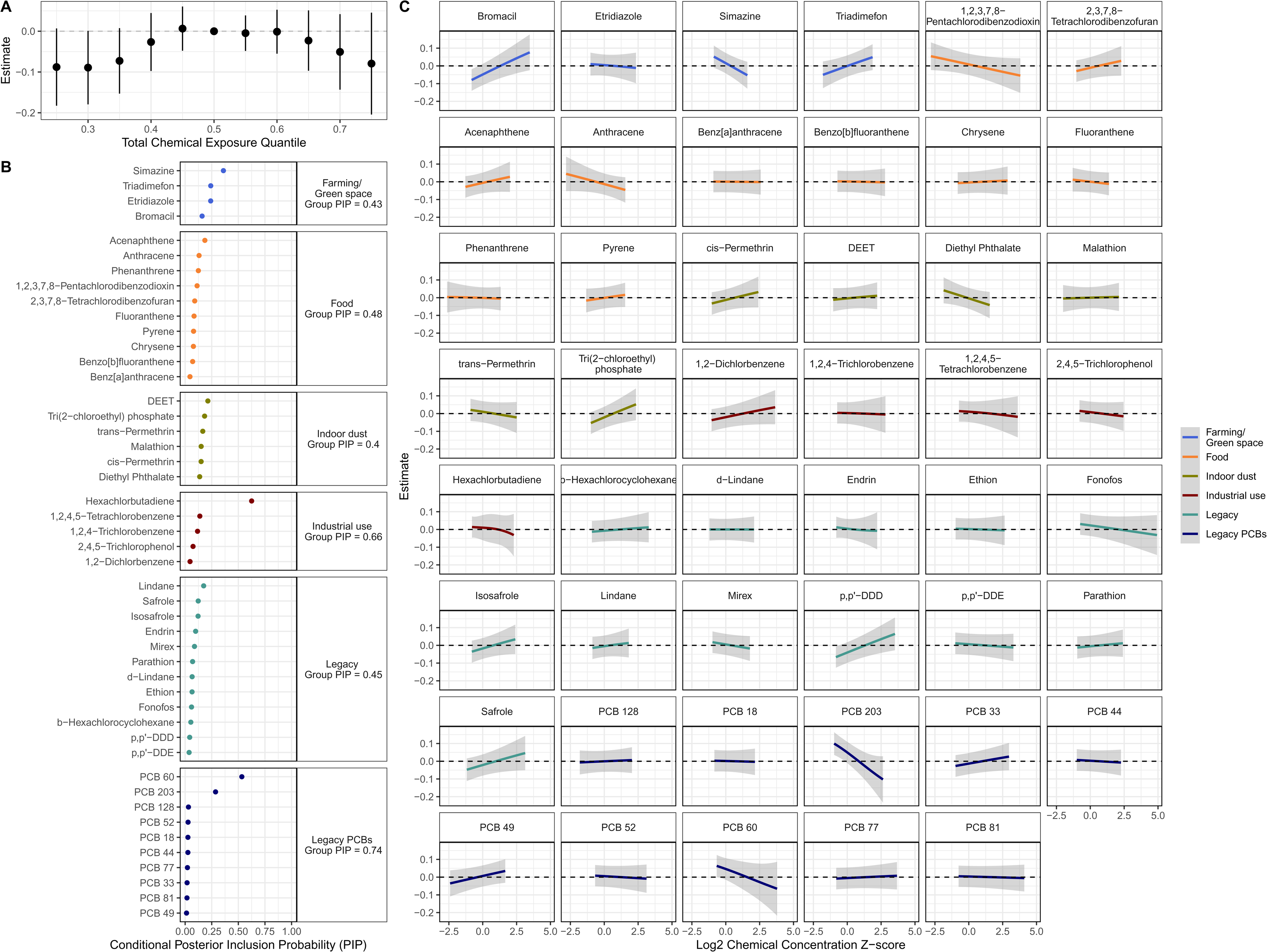
Organic pollutant mixture effect on global cognitive score. In A, the joint estimate of the relationship between the mixture all organic pollutants and the global cognitive score when each pollutant was fixed between the 25^th^ and 75^th^ percentile, with 5% increments, when each of the pollutants was fixed at their median as the reference. In B, the posterior inclusion probabilities (PIP) for the pollutant groups (indicated in the facet label) and each pollutant. The PIP gives an estimate of the number of times a group or pollutant was included in the model, providing a means to assess the importance of the group or pollutant (higher PIP indicating greater importance). In C, the univariate relationship between each pollutant and the global cognitive score, while fixing all other pollutants at their median. The grey shaded area around the lines shows the 95% credible intervals.

### 3.6 Mixture analysis of association between chemicals and cognitive domains

The estimated overall mixture effects for the four cognitive domains are shown in Figure S3. There was no dose-response relationship between the pollutant mixture and episodic memory. We estimated non-linear overall mixture effects for vocabulary and fluid reasoning. Overall, for vocabulary, the estimates decrease and are significant at higher percentiles and for fluid reasoning, the estimates increase and are significant at lower percentiles. A positive overall mixture effect was estimated for perceptual speed that was statistically significant at lower percentiles.

The group and conditional PIPs when estimating each individual cognitive domain are shown in Figure S4. Legacy PCBs had a high PIP for all cognitive domains, pollutants of industrial use had high PIP for perceptual speed, fluid reasoning, and vocabulary, and chemicals in food had high PIP for perceptual speed. The pollutant-specific independent exposure-response curves can be found in Supplemental Figures 5-8. Some relationships of note (and their direction of association in parentheses) include: for episodic memory PCB 60 (negative), 1,2-dichlorobenzene (positive), and 1,2,3,7,8-pentachlorodibenzodioxin (negative); for fluid reasoning: bromacil (positive), simazine (negative), triadimefon (positive), trans-Permethrin (negative), tri(2-chloroethyl)phosphate (positive), PCB 203 (negative), PCB 52 (negative); for perceptual speed: bromacil (positive), triadimefon (positive), acenaphthene (positive), cis-Permethrin (positive), diethyl phthalate (negative), b-hexachlorocyclohexane (positive), mirex (negative), PCB 203 (negative), PCB 33 (positive); and for vocabulary: bromacil (positive), anthracene (negative), cis-Permethrin (negative), malathion (positive), hexachlorbutadiene (negative), safrole (positive), PCB 60 (negative) and PCB 81 (negative).

Upon inspection of trace plots, those for residual variance suggested poor mixing for the episodic memory and fluid reasoning model. This remained the case even after increasing the number of MCMC chains.

## 4. Discussion

This is the first study to show a relationship between exposure to an environmental mixture of organic pollutants and cognitive domains that capture cognitive aging in a cohort of cognitively normal adults. The overall estimate of the relationship between the final mixture of 49 pollutants and a composite score of cognitive aging showed an inverted U-shaped relationship. Legacy PCBs were identified as the most important group contributing to this association, followed by chemicals of industrial use. Within legacy PCBs, PCB 60 and PCB 203 were the highest contributors. Both had a negative relationship with the global cognitive score, when controlling for all other pollutants. Although associations with other chemicals were null, we found some suggestive evidence of an association with some of the pollutants (e.g., bromacil and 1,2,3,7,8-pentachlorodibenzodioxin), but the direction of these associations was not consistent within their source groups or across these pollutants.

Exposure to PCBs has previously been studied in a few different ways: as individual congeners [15,34], as the total sum of all PCBs [34,35], commercial PCB mixture [36], or sum of PCBs classified as low-vs. high-chlorinated [13,14]. Exposure to PCBs has been associated with poor cognitive function across adulthood [14,37] and in older adults [13,15,34–36,38]. They have also been negatively associated with neurodevelopment [39,40]. Individual congeners that have been previously associated with cognitive function include PCB 146, PCB 105, PCB 118, PCB 138, PCB 170, PCB 180, and PCB 194 [15,34]. A study that applied a mixtures approach found that PCBs (both low- and high-chlorinated) were more important contributors than organochlorine pesticides in the negative relationship observed with cognitive function [13,14]. Another study applied path analysis to determine the relationship between PCB congeners and cognitive function among older adults in an NHANES population (1999-2002). After controlling for co-exposure to known neurotoxic metals, they found a negative relationship between PCB 146 and cognitive function [15]. While dioxin-like PCBs, which bind to the aryl hydrocarbon receptor, have been considered to have greater toxicity than non-dioxin-like PCBs, evidence from both epidemiological and toxicological studies suggest non-dioxin like PCBs have greater neurotoxicity [15,41].

Mechanistic studies have shown that exposure to PCBs reduced long-term potentiation in CA1 hippocampal neurons, as well as synaptic transmission in rats [42,43]. Exposure to PCBs has also been shown to reduce cellular levels of dopamine and its metabolites in the caudate, putamen, substantia nigra, and hypothalamus [44]. While dioxin-like PCBs have been identified as drivers of hepatotoxicity and immunotoxicity, there is poor evidence that they play a role in dopamine-related toxicity ascribed to PCBs. The reduced cellular dopamine content is a result of interaction at specific molecular sites that prefer ortho, or ortho-, para-substituted PCB congeners, which are non-dioxin like PCBs [41]. Apart from altered dopamine content, PCB mixtures have also been shown to alter dopamine transporter levels in the striatum [45]. Studies have also shown that PCB exposure alters calcium signaling, possibly through ryanodine receptors in nerve cells [46,47], which may have consequences for neurodevelopment and neurodegeneration. Indeed, PCBs have been related to Parkinson’s disease pathology [48]. Other potential mechanisms through which PCBs may induce neurotoxicity include mitochondrial damage, generation of reactive oxygen species, and increased membrane fluidity [46,49].

PCBs have been phased out and banned from use in sealants and coolants. However, they are highly lipophilic and persist in the environment. Exposure to PCBs mainly occurs through contaminated food: namely fish, seafood, and dairy. Inhalation is also an important route of exposure to low-chlorinated PCBs since these are more readily volatilized, which is of particular concern in occupational settings [50]. There is also mounting evidence that there may be inadvertent formation of PCBs (non-legacy PCBs), during manufacture of paint pigments [51,52]. Decline in levels of PCBs in the environment were seen soon after their ban, but their levels have since stabilized. The Stockholm convention has outlined an action plan to cease all use of PCBs by 2025 and make efforts for environmentally sound management of waste that contains PCB by 2028 [53,54]. It remains to be seen if these actions will reduce the threat posed by PCBs to the public’s health, like that identified through this study.

The overall association between exposure to the chemical mixture we examined, and the global cognitive score showed an inverted U-shaped relationship. In toxicology, such a relationship suggests a hormetic dose response, i.e., stimulation at low dose and inhibition at high dose [55–57]. The observed relationship could also be explained by a maximal adaptability model (MAM), which has previously been used to describe the inverted-U or extended-U shaped relationship between cognitive function and temperature [3,58]. However, the MAM model has not been shown to adequately explain the effect of chronic exposure to environmental chemicals that may be stored in different physiological compartments, including the brain. While the overall effect estimates below the median increased with increasing percentiles of the chemical levels, the estimates were negative, suggesting that the stimulatory effect was not accompanied with enhanced function, as is usually expected with a hormetic effect. Therefore, neither model adequately explain the observed dose-response relationship.

Our study has several novel aspects; however, some limitations have been identified. Given the large number of pollutants, this study was underpowered to find small or moderate effects. Others have shown age and sex-specific effects on the relationship between PCBs and cognitive function [14,36], but we were underpowered to conduct an analysis stratified by sex and age. While we captured many chemical exposures, we were unable to account for co-exposures that were not measured. In the future, we plan to use non-targeted high-resolution mass spectrometry-based exposomic data to further explore the role of other co-exposures. The serum samples collected were not from a fasting state. Since levels of organochlorine chemicals measured in serum collected in non-fasting state tend to be higher [59], the concentrations reported in our study may be higher than levels measured in fasting samples. Additionally, the results may not be generalizable since the study population was highly educated and met strict inclusion criteria. Finally, there is the possibility of residual confounding that we did not adequately address. Despite these limitations, our study has several strengths: 1) we measured chemical exposure at two time points ∼ 5 years apart, better capturing variation in exposure; 2) we included participants from a wide age range; 3) we estimated mixture effects of pollutants from different classes and environmental sources; and 4) we applied a mixtures analysis approach that allowed us to consider non-linear relationships between exposure and outcome.

In conclusion, to our knowledge, this is the first study to identify an association between exposure to a mixture of organic pollutants and cognitive domains that capture key aspects of cognitive aging in a cohort of cognitively normal adults. PCBs 60 and 203 were the most important pollutants in the mixture and were both negatively associated with the global cognitive score. PCBs have been shown to induce neurotoxicity, but mechanistic studies are needed to understand the effect of these two specific PCB congeners. Further, steps should be taken to reduce the public’s exposure to PCBs, which while banned, remain a significant threat to health, especially in an aging population.

## Data Availability

All data produced in the present study are available upon reasonable request to the authors.

https://github.com/vrindakalia/chemicals_and_reference_abilities

## Acknowledgements

We would like to thank the participants of the Cognitive Reserve and Reference Ability Neural Network study.

## Conflicts of Interest

The authors declare that the research was conducted in the absence of any commercial or financial relationships that could be construed as a potential conflict of interest.

## Consent Statement

The patients/participants provided their written informed consent to participate in this study.

## Source of Funding

The analysis in this manuscript was supported by the Pilot Projects Program from the Center for Environmental Health in Northern Manhattan funded by National Institute of Environmental Health Sciences (NIEHS) under grant P30 ES009089. VK was supported by National Institute on Aging (NIA) grant P20 AG093975. MAK was supported by NIEHS grants R01 ES028805 and P30 ES009089, and NIA grant P20 AG093975. GWM was supported by NIEHS grants R01 ES023839 and U24 ES036819, and NIA grant P20 AG093975. YG was supported by NIA grant R01 AG061008.

## Notes

### Competing Interest Statement

The authors have declared no competing interest.

### Author Declarations

The IRB of Columbia University gave ethical approval for this work.

