## Supplementary figures for "Mixture of organic pollutants is associated with cognitive aging"

Corresponding author:

Vrinda Kalia, PhD, MPH

650 W 168^th^ street; Room-1618

New York, NY 10032

**Supplementary Figures**

**Figure S1.** The distribution of correlation coefficients between each pollutant group, assigned based on use/source. The bars to the right of the dotted red line show the distribution of positive correlation coefficients.

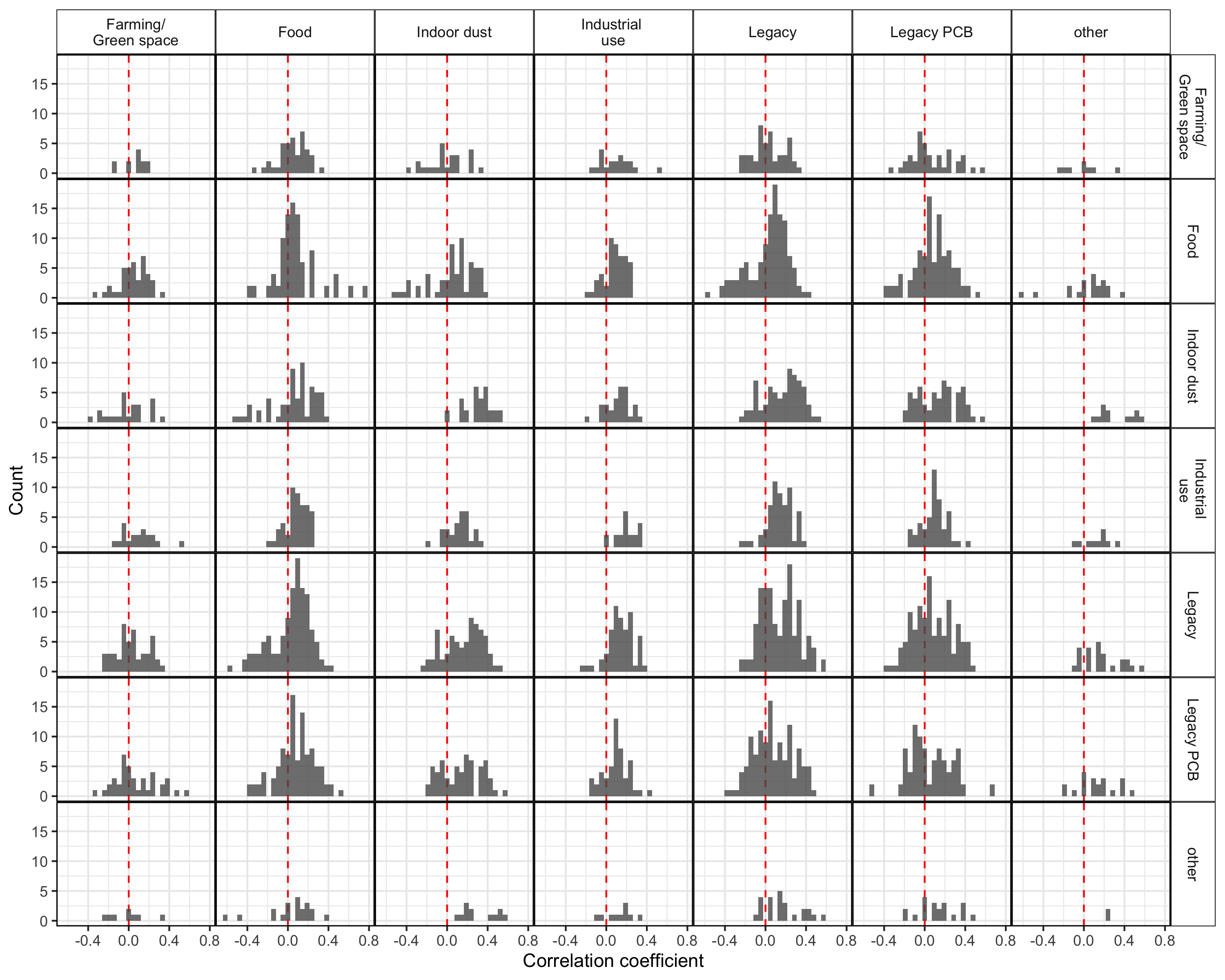

**Figure S2. Individual pollutant models.** The forest plot shows the generalized estimating equation derived effect estimate and 95% confidence interval, calculated using robust standard errors, of each pollutant and the four cognitive domains as well as the global score. The models were adjusted for age, sex, race/ethnicity, APOE-ε4 allele status, and batch of analysis.

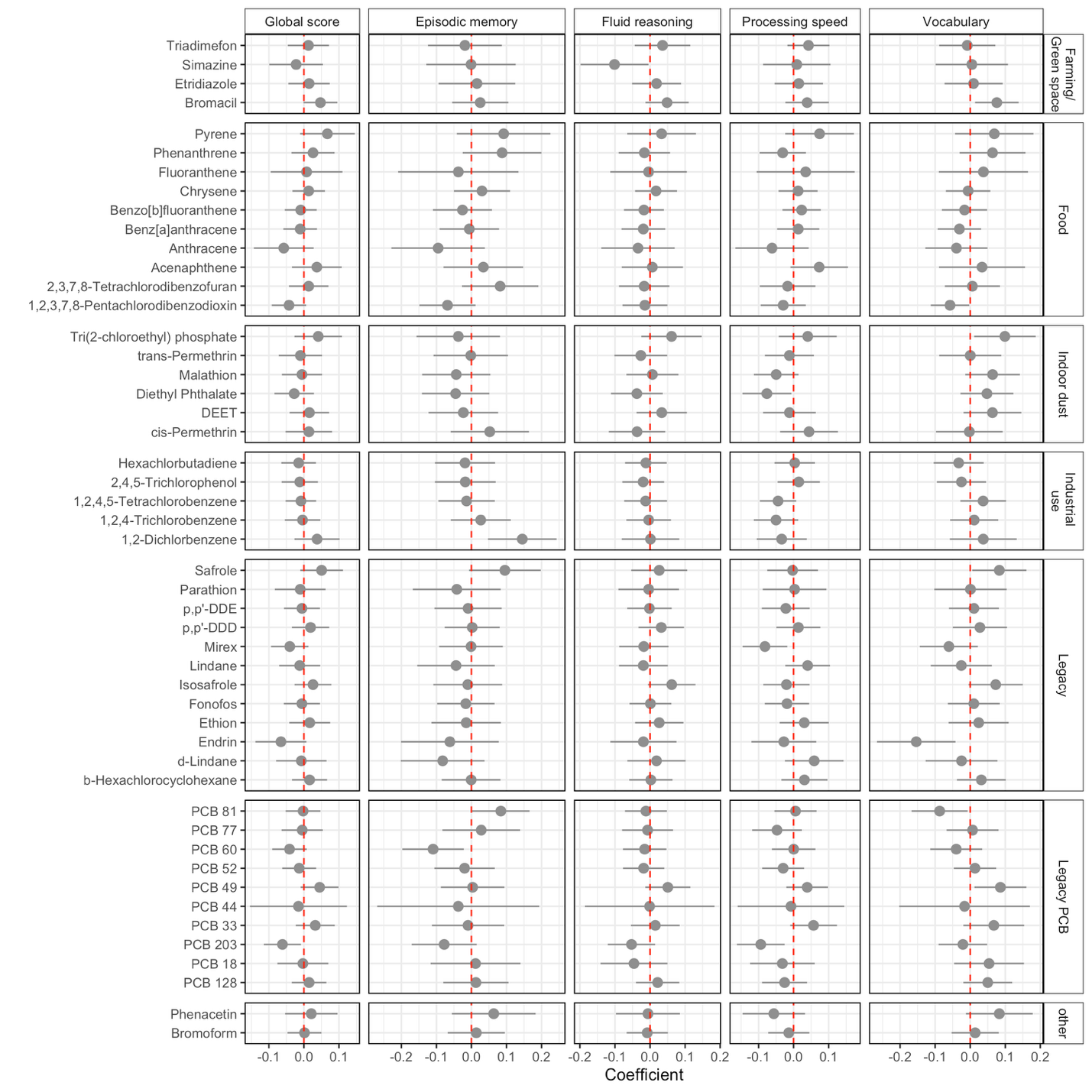

**Figure S3.** The joint estimate of the relationship between the mixture of all organic pollutants and each of the cognitive domains when each pollutant was fixed between the 25^th^ and 75^th^ percentile, with 5% increments, when each of the pollutants were fixed at their median as the reference.

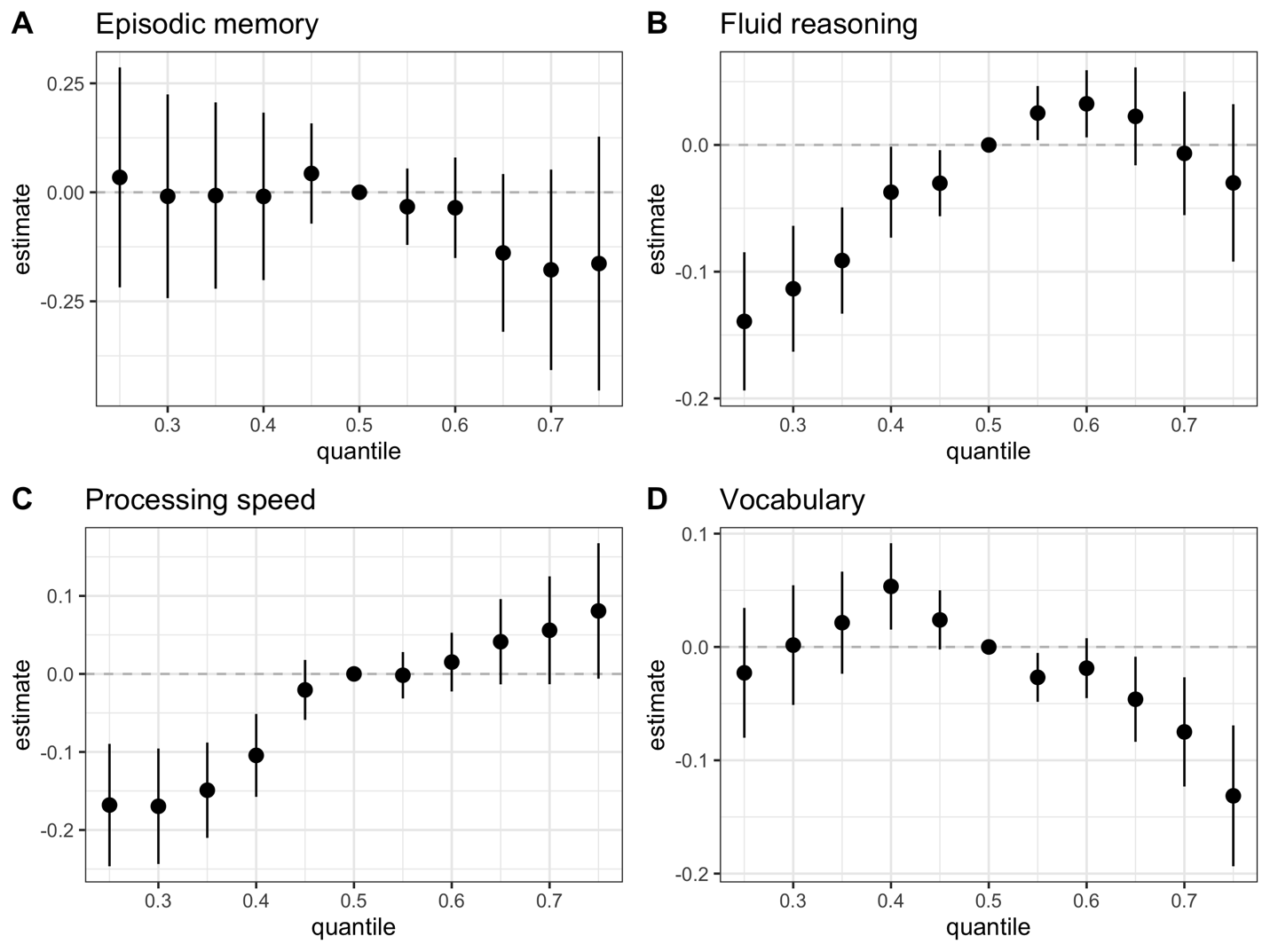

**Figure S4.** **Pollutants of importance in estimating the relationship between the mixture and each cognitive domain.** The group PIP and individual (or conditional) PIP (indicated by shape, the square referring to group PIP) are shown for each outcome (indicated by color).

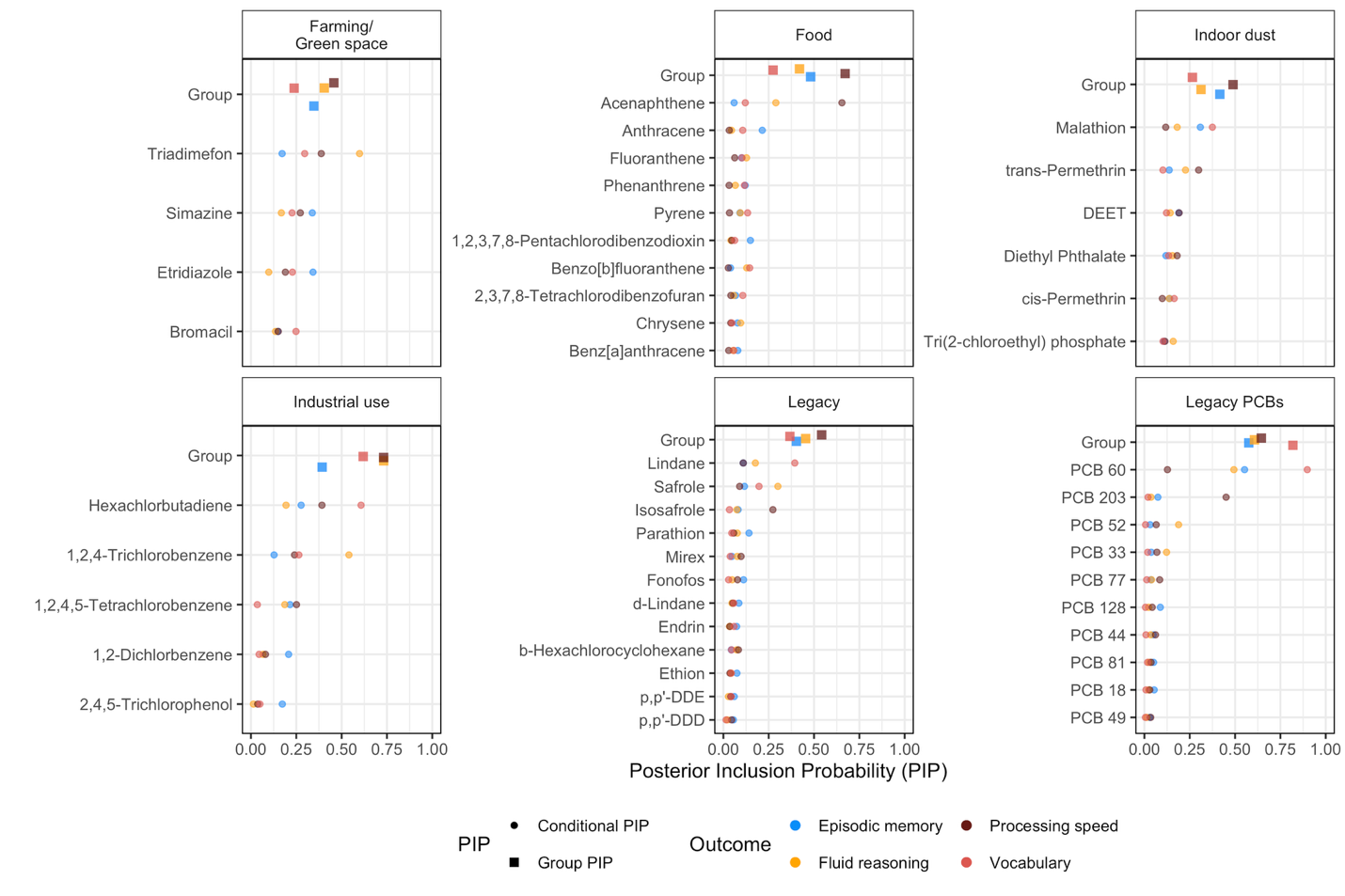

**Figure S5**. Episodic memory. The univariate relationship between each organic pollutant and the episodic memory cognitive domain, while fixing all other chemicals at their median. The grey shaded area around the lines shows the 95% credible intervals.

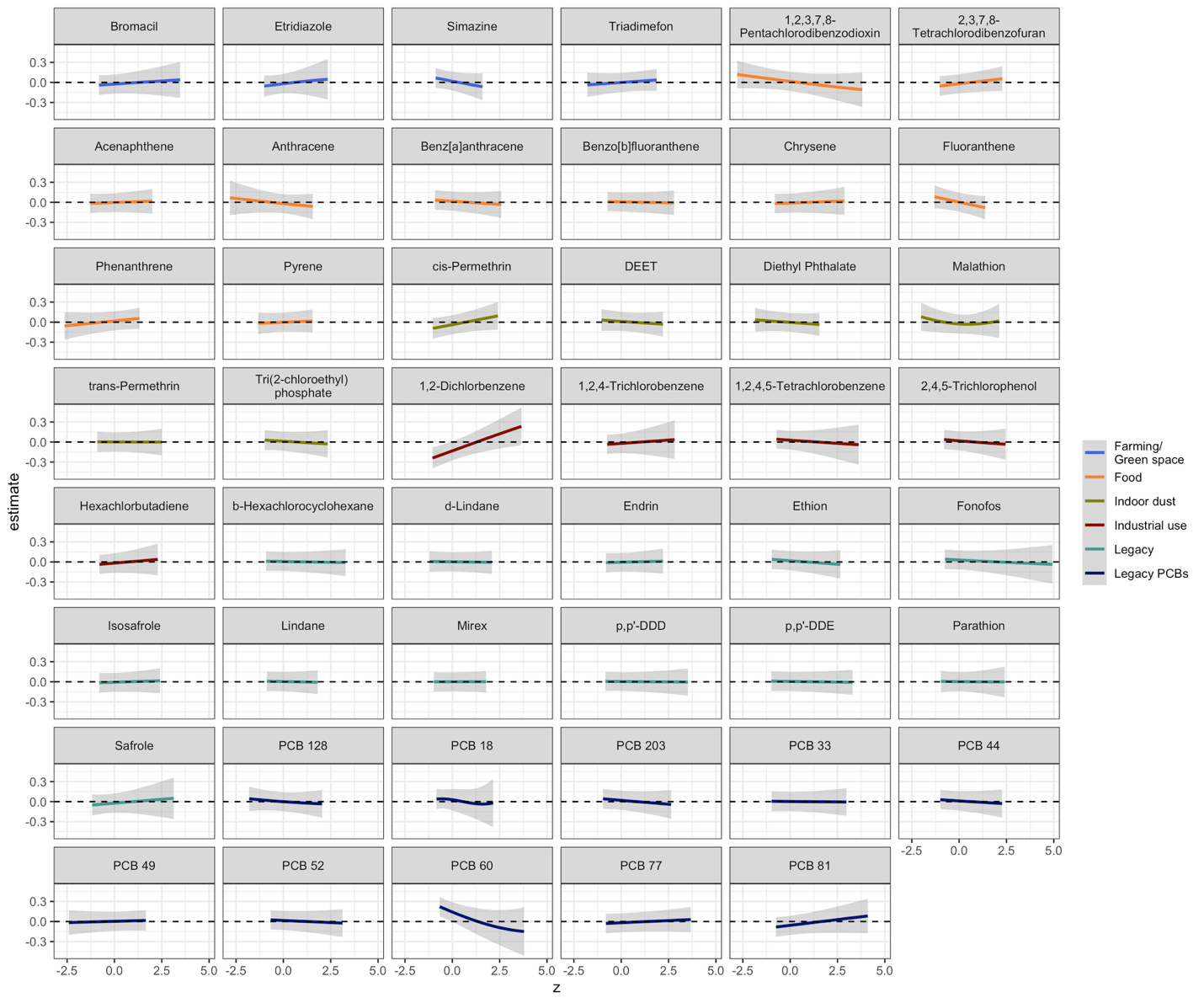

**Figure S6**. Fluid reasoning. The univariate relationship between each organic pollutant and the fluid reasoning cognitive domain, while fixing all other chemicals at their median. The grey shaded area around the lines shows the 95% credible intervals.

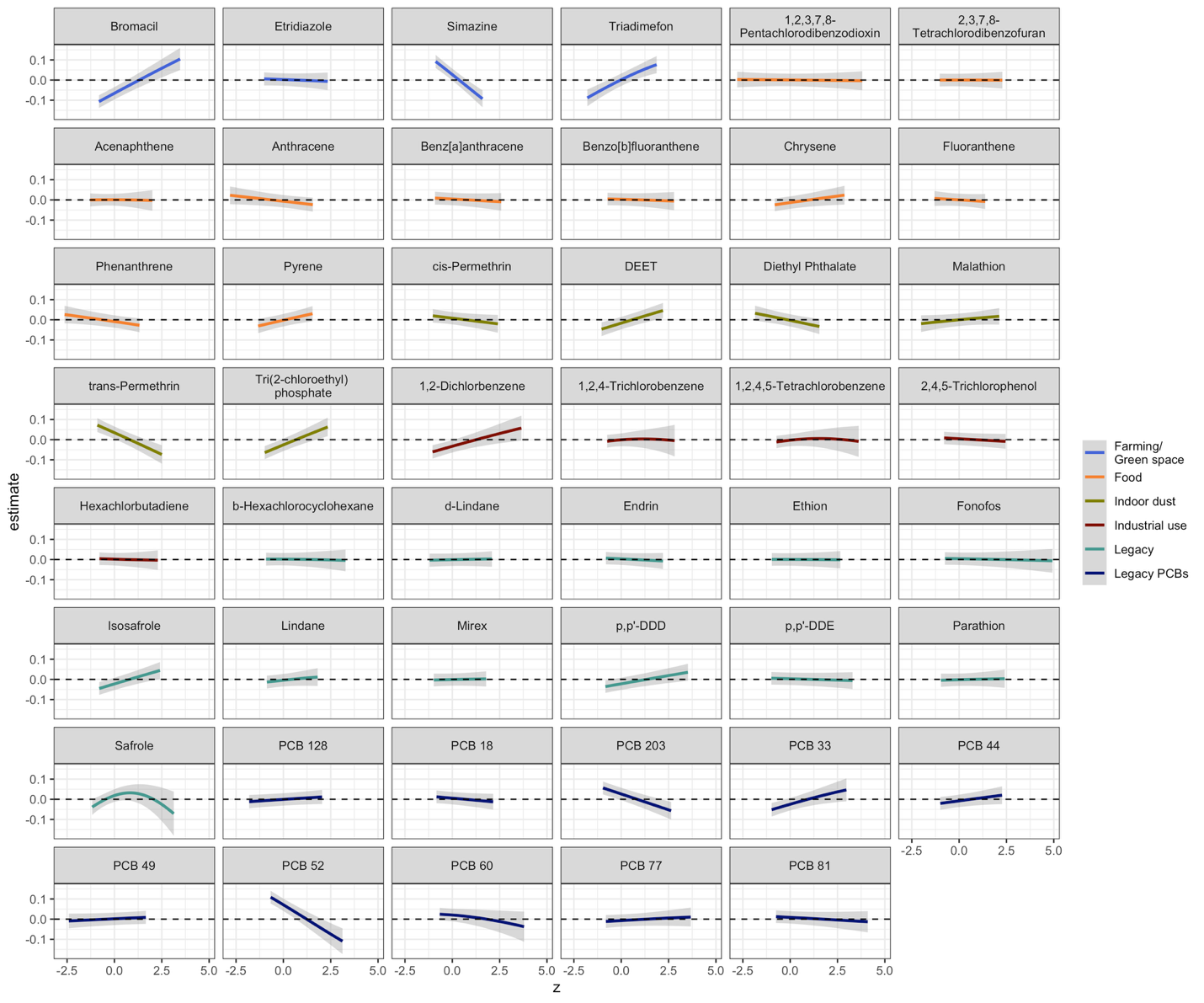

**Figure S7**. Processing speed. The univariate relationship between each organic pollutant and the perceptual speed cognitive domain while fixing all other chemicals at their median. The grey shaded area around the lines shows the 95% credible intervals.

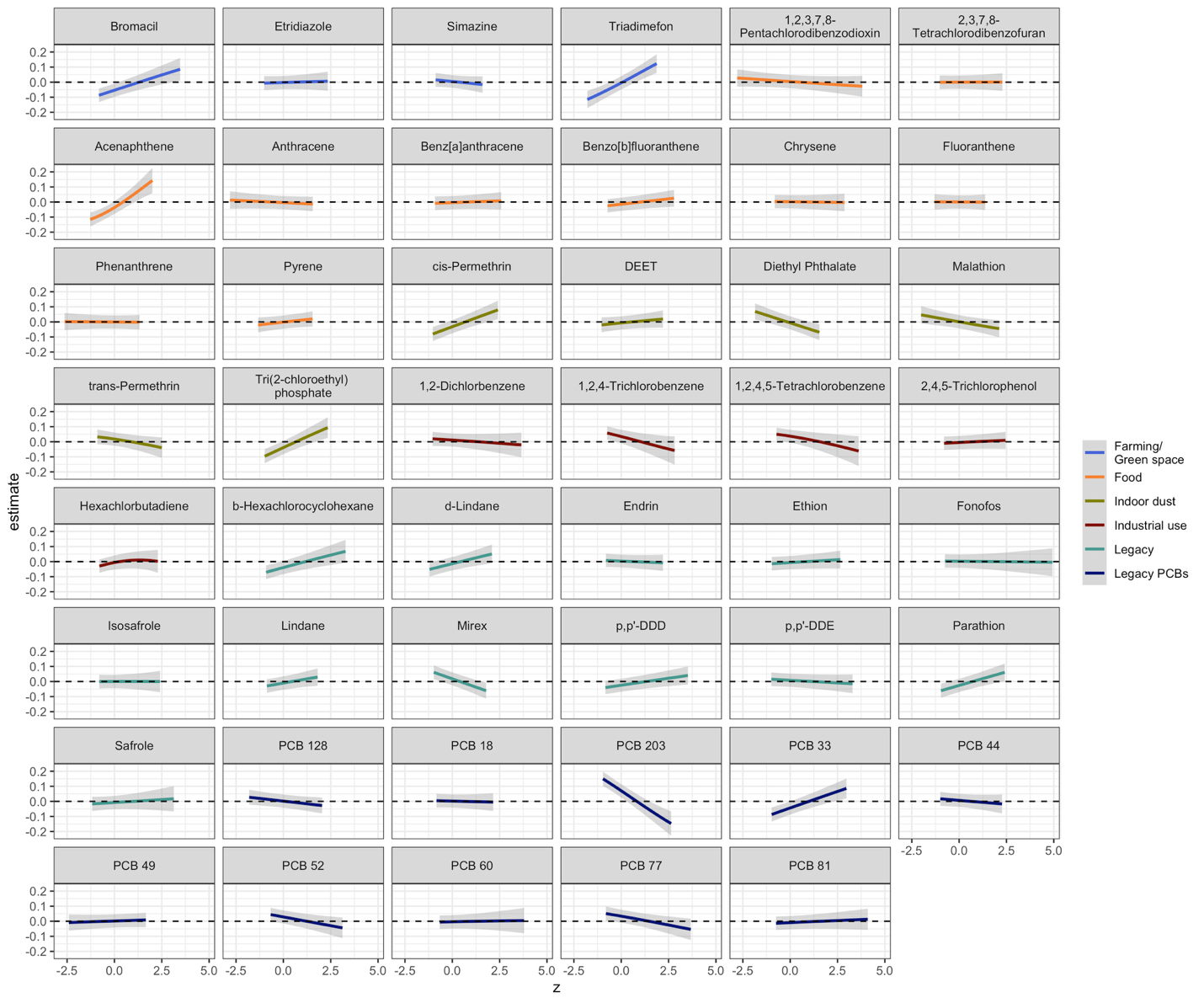

**Figure S8**. Vocabulary. The univariate relationship between each organic pollutant and the vocabulary cognitive domain, while fixing all other chemicals at their median. The grey shaded area around the lines shows the 95% credible intervals.

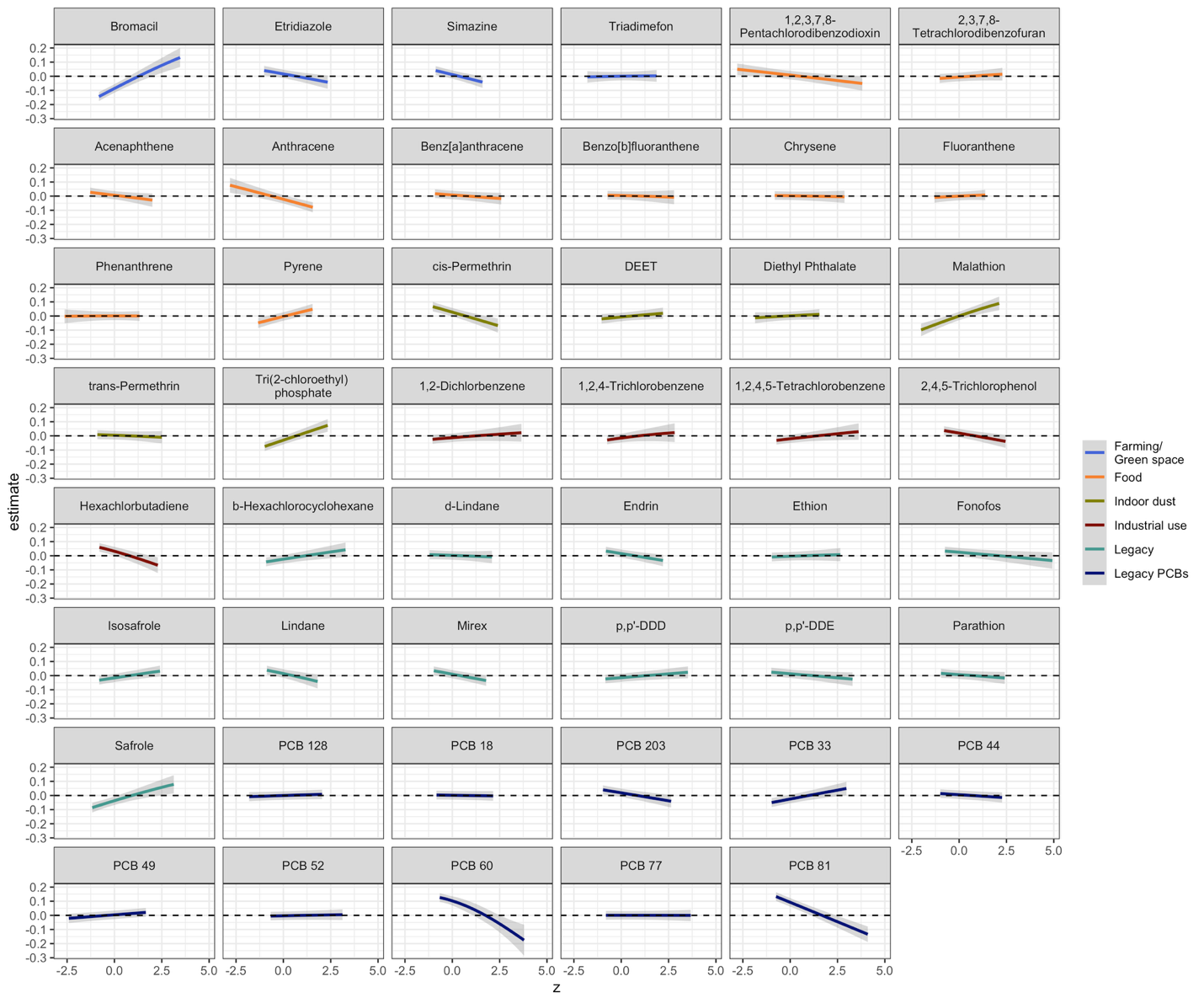
